# Ultra-low coverage fragmentomic model of cell-free DNA for cancer detection based on whole-exome regions

**DOI:** 10.1101/2024.02.06.24302178

**Authors:** Apiwat Sangphukieo, Pitiporn Noisagul, Patcharawadee Thongkumkoon, Parunya Chaiyawat

**Affiliations:** Center of Multidisciplinary Technology for Advanced Medicine (CMUTEAM), Faculty of Medicine, Chiang Mai University, Chiang Mai, Thailand

## Abstract

Cell-free DNA (cfDNA) has shown promise as a non-invasive biomarker for cancer screening and monitoring. The current advanced machine learning (ML) model, known as DNA evaluation of fragments for early interception (DELFI), utilizes the short and long fragmentation pattern of cfDNA and has demonstrated exceptional performance. However, the application of cfDNA-based model can be limited by the high cost of whole-genome sequencing (WGS). In this study, we present a novel ML model for cancer detection that utilizes cfDNA profiles generated from all protein-coding genes in the genome (exome) with only 0.08X of WGS coverage. Our model was trained on a dataset of 721 cfDNA profiles, comprising 426 cancer patients and 295 healthy individuals. Performance evaluation using a ten-fold cross-validation approach demonstrated that the new ML model using whole-exome regions, called xDELFI, can achieve high accuracy in cancer detection (Area under the ROC curve; AUC=0.896, 95%CI = 0.878 - 0.916), comparable to the model using WGS (AUC=0.920, 95%CI = 0.901 – 0.936). Notably, we observed distinct fragmentation patterns between exonic regions and the whole-genome, suggesting unique genomic features within exonic regions. Furthermore, we demonstrate the potential benefits of combining mutation detection in cfDNA with xDELFI, which enhance the model sensitivity. Our proof-of-principle study indicates that the fragmentomic ML model based solely on whole-exome regions retains its predictive capability. With the ultra-low sequencing coverage of the new model, it could potentially improve the accessibility of cfDNA-based cancer diagnosis and aid in early detection and treatment of cancer.

## Introduction

Cell-free DNA (cfDNA) is a term for DNA fragments that are mostly released into body fluids from various sources, such as apoptotic or necrotic cells, as well as from active secretion [1]. The elevation of cfDNA in the blood can be an indicator of various health conditions, especially cancer [2, 3]. Analysis of cfDNA in the context of a liquid biopsy is gaining popularity in the field of oncology as it shows variety of benefits, for example, non-invasiveness that require only blood draw, the ability of each sample to reflect all tumor lesions in the body, and ability to real-time monitor cancer progression due to short half-life of cfDNA (∼2 h) [4]. A particular subset of cfDNA known as circulating tumor DNA (ctDNA), which contains genetic alterations associated with cancer development, holds immense potential for diagnosing the disease, tracking cancer progression, evaluating treatment response, and identifying potential therapeutic targets [1]. However, the extremely low concentration of ctDNA in plasma, which is lower than 0.01% of the total cfDNA concentration [5], presents a significant obstacle for accurate detection and analysis.

Numerous studies demonstrated that the fragment length of cfDNA derived from normal cells is about 166 bp on average, while the cfDNA derived from tumor cells is more fragmented with the size between 90–150 bp [6–9]. Furthermore, there has been a notable observation of a positive correlation between the proportion of short cfDNA fragments (<150 bp) and the tumor DNA fraction present in the plasma [7]. In addition, numerous research studies have examined the possibility of using shorter cfDNA fragments to improve the detection of copy number variations (CNV) and single nucleotide variations (SNV) in the cancer patients [7, 10, 11]. In recent years, there has been a growing interest in the study of cfDNA fragmentation patterns, known as fragmentomics [12, 13], including various aspects of cfDNA fragmentation, such as fragment sizes, abundance, integrity, end motifs, window protection score, and preferable end coordinates [12, 14]. The advancement of next-generation sequencing technology has enabled whole-genome sequencing (WGS) showing that cfDNA fragment size distribution patterns in cancer patients are more variable than those in healthy individuals [15]. These differences in cfDNA fragmentation patterns reflect changes in chromatin structures, as well as other genomic and epigenomic abnormalities in cancer [16] providing a framework for developing diagnostic tools [17]. Recently, a machine-learning (ML) approach was applied to learn the pattern of cfDNA fragmentation from low-coverage WGS data, known as DNA evaluation of fragments for early interception (DELFI), showing excellent performance in the classification of cancer-carried individuals and healthy individuals [15]. DELFI evaluated the fragment size coverage of short cfDNA fragments (100–150 bp) and long cfDNA fragments (151–220 bp) inside 100 kilobase (Kb) non-overlapping consecutive bins and integrated them into a 5-megabase (Mb) non-overlapping consecutive window. The aberrations of the ratio between short cfDNA fragments and long cfDNA fragments within each window is increased in cancer patients, but not in healthy individuals. This cfDNA fragmentation size difference was utilized as a key feature in gradient-boosting model and showed outstanding performance with an overall area under the curve (AOU) of 0.94 [15]. The performance of DELFI model has been further improved by reducing feature dimension, applying different feature extraction strategy, and utilizing ensemble algorithm [13, 18]. Nevertheless, its practical application is limited by the current WGS cost. While reducing the sequencing coverage to 0.1X WGS has been demonstrated as an effective cost-saving measure for estimating tumor fraction [19], it also introduced significant alterations in the fragmentation profiles [13, 15], which could potentially impact the accuracy and reliability of DELFI score.

While most of the human genome is non-coding and its function remains largely unknown, only 1% of the genome is comprised of protein-coding regions, known as exons. Exons are the protein-coding regions of a gene that are transcribed into mRNA and ultimately translated into proteins. The sequences of exons are typically highly conserved across different species, reflecting the importance of these regions in protein function and evolution [20]. Mutations in exons can lead to altered protein function and are often associated with genetic diseases. With the availability of large databases of known SNPs and known pathogenic variants, whole-exome sequencing (WES), an alternative approach to whole-genome sequencing (WGS) by targeting the exonic regions, have been extensively applied to identify casual mutations in cancer patients with much lower cost than WGS [21]. Recently, researchers have been exploring the use of cfDNA fragmentation profiles at exonic regions to infer gene expression, which can apply to various clinical applications such as tumor detection, subtype classification, treatment response assessment, and prognostic implications [22].

In this study, we present a novel approach for developing the DELFI model that can classify cancer patients and healthy individuals based on exonic regions. Our new exome-based cfDNA fragmentation model, called xDELFI, can efficiently distinguish between cancer patients and healthy individuals and can classify the tissue of origin with reasonable accuracy. Furthermore, our study showed that combining xDELFI with mutation information can further enhance the prediction performance, which highlights the potential benefits of utilizing WES data in mutation calling and xDELFI score prediction. The new model paves the way to create more cost-effective methods for cancer diagnosis and monitoring.

## Methods

### Data collection

The paired-end cfDNA whole-genome sequencing samples from four different articles stored in FinaleDB database were collected. In total of 426 samples from 16 cancer types and 295 healthy individuals were processed [7, 15, 16, 23] (Supplementary Figure 1). The data was pre-processed steps, which had been proceed in finaleDB, to have high quality data by 1) trimming all sequences to 50 bp to minimize possible batch effects, 2) exclude un-properly mapped in pair, 3) exclude non-primary alignments, 4) exclude reads with mapping quality < 30, and 4) remove duplicated reads. Data quality control was proceeded by measuring correlation of the whole genome sequence coverage observed in this study and those reported in the original article (Supplementary Figure S3).

### DELFI and exomeDELFI score calculation

DELFI score calculation is followed [15]. The original script of DELFI is available in https://github.com/cancer-genomics/delfi_scripts. Briefly, all chromosomes have been split into consecutive, non-overlapping 100 kb bins. The lowest coverage bins (top10%) and fragments falling in the Duke blacklisted regions (http://hgdownload.cse.ucsc.edu/goldenpath/hg19/encodeDCC/wgEncodeMapability/) were removed. Short (100 and 150 bp) and long (151 and 220 bp) cfDNA fragment coverage of each 100k bin were counted. For exomeDELFI, only fragments falling in the exome regions based on Agilent - SureSelect All Exon V7 (bed file is available at https://github.com/mobidic/BARMEN) were counted. Loess regression-based approach is applied to account for GC content bias for each 100kb bin in the genome of each sample. The 100k bins were combined into 5Mb bins (504 bins). Therefore, the total number of bins/features from short and long fragment coverage is 1,008. Then, GC-adjusted short and long fragment coverage were centered and scaled for each sample to have mean 0 and unit standard deviation. Feature selection, performed only on the training data (in each cross-validation run), removed bins that were highly correlated (correlation > 0.9) or had near zero variance. Gradient boosting machine (GBM) was implemented using the caret package in R with parameters of n.trees=150, interaction.depth=3, shrinkage=0.1, and n.minobsinside=10. The prediction error was evaluated by performing ten-fold cross validation (CV) repeated ten times. The final DELFI score is an average probability of the cancer class from ten repeated CV.

### xDELFI score calculation

Firstly, we applied three fragment size thresholds, which are short (100-150), medium (150-220), and long (>220), to count the number of fragments in each 100k bin along the genome, instead of using short (100-150) and long (150-220) threshold in previous study (Figure 1B). In addition to fragment length coverage, overall fragment coverage is counted by summation of all fragments in each 100k bin. Secondly, loess regression-based approach is applied to account for GC content bias for each 100kb bin. Thirdly, the 100k bins were combined into 5Mb bins (504 bins) for each chromosome arm. Then, the fragment size distribution (FSD) of 5bp bin in range of 100 to 220bp (24 bins) in each chromosome arm (39 arms) was calculated, and loess regression-based approach is applied to account for GC content bias for each bin. Therefore, the total number of features from fragment length coverage (504×3), overall coverage (504), and FSD (24×39) is 2,952. Feature selection method was applied only on the training data to remove highly correlated features (correlation > 0.9) or uninformative features (zero variance). Finally, two different machine learning models, GBM and support vector machine (SVM), were combined as stacking ensemble by GBM to train xDELFI model. The stacking ensemble model was implemented using caretEnsemble package with GBM parameters of n.trees=70, interaction.depth=3, shrinkage=0.1, and n.minobsinside=10. Ten-fold CV repeated ten times was conducted to evaluate the prediction error and the average probability of the cancer class was used as the final xDELFI score. xDELFI script is freely available at Github: https://github.com/asangphukieo/xDELFI.

**Figure 1.**
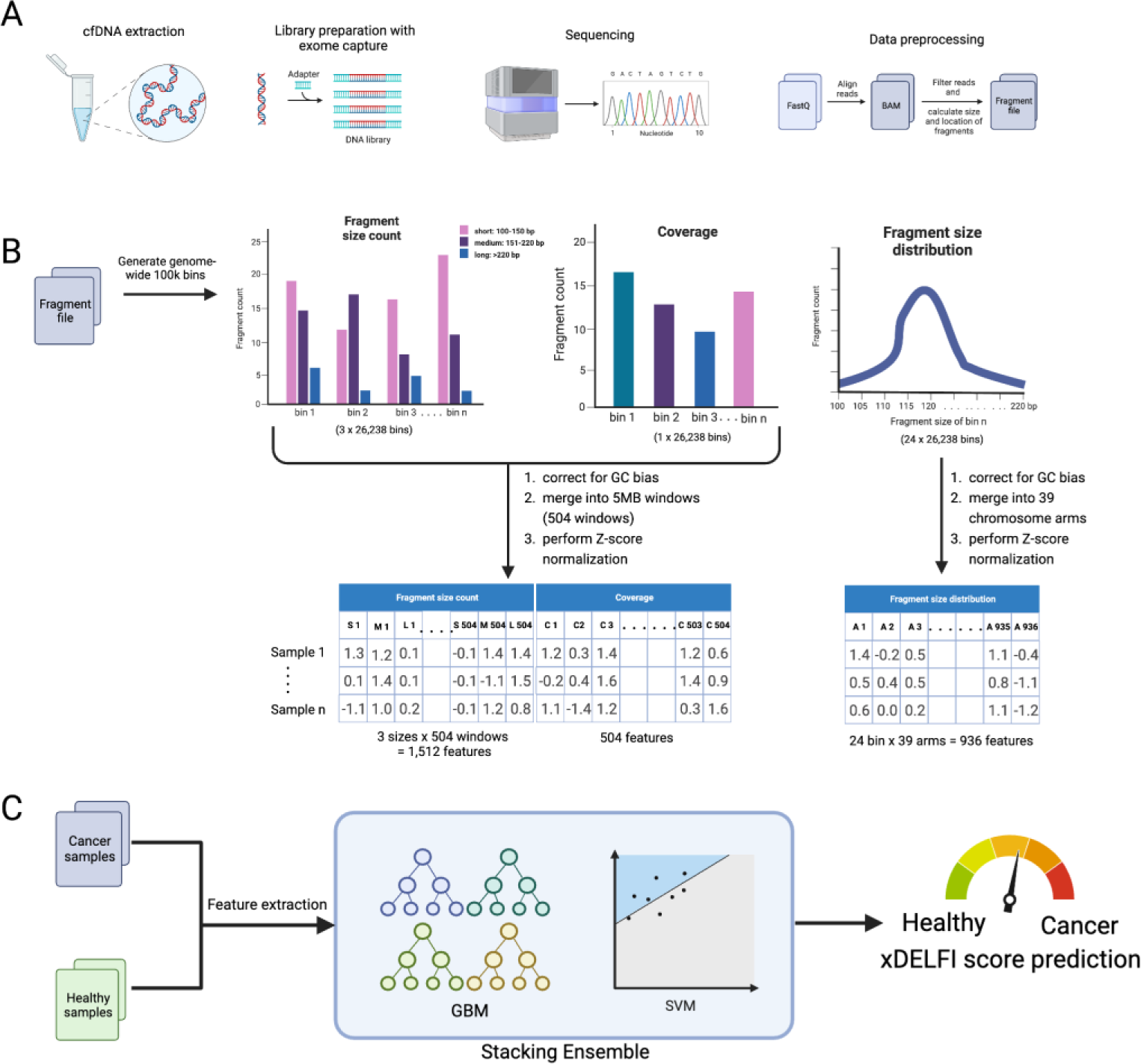
Schema of xDELFI calculation (A) general procedure of cell-free DNA process consists of blood collection, cfDNA extraction, library preparation for whole-genome sequencing or whole-exome sequencing, sequencing using next-generation sequencing platforms, and data preprocessing. (B) xDELFI feature extraction contains three fragment size count (100-150bp, 151-220bp, and >220bp), overall fragment count in each 100k bin along the genome, and fragment size distribution of 5bp bin in in each chromosome arm. Loess regression-based approach is applied to correct for GC bias and z-score is applied for normalization. (C) Gradient boosting machine (GBM) is combined with support vector machine (SVM) algorithms by stacking ensemble method to learn normalized fragmentation pattern and generate xDELFI score.

## Results

### DELFI score based on exonic regions show good performance for cancer detection

To develop the whole exome-based DELFI model, the paired-end cfDNA whole-genome sequencing samples from four different articles stored in FinaleDB database were used. In total of 426 samples from 16 cancer types and 295 healthy individuals were processed (Supplementary Figure S1). Initially, we examine the reproducibility of the original DELFI score using short and long fragment coverage as a model feature. Reconstructing DELFI model with the same parameters employed in Cristiano et al., 2019 of the present dataset was conducted and showed high prediction performance with AUC score of 0.920 (95%CI = 0.901 – 0.936). The DELFI scores have a strong Pearson correlation coefficient (*r*) with those published in the original publication (*r* = 0.855) (Supplementary Figure S2), demonstrating high reproducibility of the DELFI score.

To observe the potential of using whole exome for DELFI score calculation, we extracted the cfDNA fragments that localized in only exon region. With the same machine learning procedure as in DELFI, we observed the good performance of DELFI score based on the whole exome, hereafter called exomeDELFI, with AUC of 0.869 (95%CI = 0.849-0.889) (Figure 2A and Figure 2B). Although the exomeDELFI performed significantly worse than DELFI model (One sample t-test *P* < 0.001), the sequence coverage for constructing the model was dramatically decreased from 2.8X whole-genome coverage to 0.08X on average (97% decrease). In addition, the trend of the score is highly correlated with DELFI score (*r* = 0.805) (Figure 2C and Supplementary Table S2). These results suggest that exome region can be used to calculate DELFI score with high prediction performance, and it paves the way in using targeted regions of whole-exome sequencing data as a source of DELFI score calculation.

**Figure 2.**
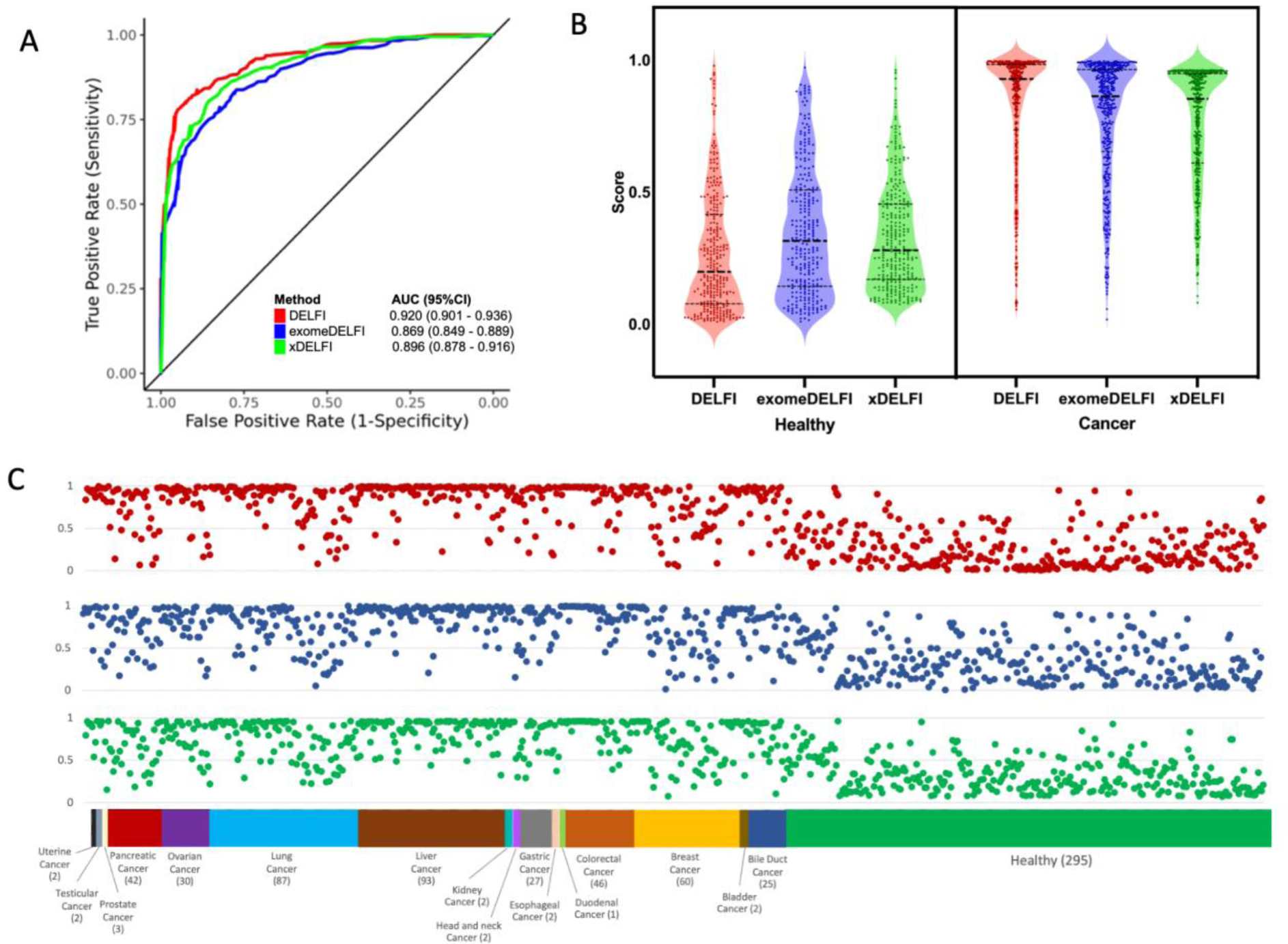
Prediction performance of DELFI score, and exomeDELFI and xDELFI visualized by (A) Receiver operating characteristic curve (ROC) and area under the curve (AUC). Violin plot (B) shows the distribution of scores across the different DELFI methods. Scatter plot (C) of the scores in different DELFI methods stratified by each cancer type. The Y-axis displays the DELFI scores, while the X-axis represents the cancer types. Red dots represent DELFI score, blue dots represent exomeDELFI and green dot represents xDELFI.

To investigate whether the fragmentomic profile from the exome regions hold distinct informative features compared to the profile from whole genome, we conducted a performance comparison between the exomeDELFI model and the DELFI model constructed using an equivalent proportion of sequencing reads. We sampled the cfDNA fragments along the genome in each sample to be equal to 0.08x whole-genome coverage and used these fragments to construct the DELFI model. Interestingly, the reduced DELFI model (AUC = 0.839; 95%CI =0.816 – 0.862) performed significantly worse than the exomeDELFI model (One sample t-test *P* < 0.001). The fragmentomic profile of short and long fragments along the genome in the reduced DELFI model was highly correlated with those used in the original DELFI model (*r* = 0.654). On the other hand, fragmentomic profile of short and long fragments used in the exomeDELFI model have no correlation with those used in the original DELFI model (*r* = - 0.030) (Table 1). These findings suggest that exomeDELFI relies on distinct genomic features differing from the features utilized by the original DELFI model to differentiate between cancer patients and healthy individuals.

**Table 1.** Correlation between whole genome-based fragmentomic profile and whole exome-based fragmentomic profile.

| Method | Source of fragmentomic profile | Whole genome coverage | AUC (95% CI) | Accuracy (95% CI) | $r^*$ |
| --- | --- | --- | --- | --- | --- |
| DELFI | Whole genome | 2.8X | 0.920<br>(0.901 - 0.936) | 0.840<br>(0.831- 0.848) | 1 |
| DELFI | Whole genome | 0.08X | 0.839<br>(0.816 - 0.862) | 0.755<br>(0.744 - 0.764) | 0.654 |
| exomeDELFI | Exome | 0.08X | 0.869<br>(0.849 - 0.889) | 0.787<br>(0.777 - 0.796) | -0.031 |

\*Pearson correlation coefficient against whole-genome fragmentomic profile

### Improvement of exome-based DELFI score

To improve the prediction performance of exome-based DELFI score, we developed a new feature extraction strategies and redesigned the model structure (Figure 1). Firstly, we have included large cfDNA fragments (>220 bp) in the calculation, as these fragments have been found in the blood and may have been released from necrotic cells [24, 25]. Thus, we applied three fragment size thresholds, which are short (100-150 bp), medium (150-220 bp), and long (>220 bp), to count for number of fragments in each 100k bin along the genome, instead of using short (100-150 bp) and long (150-220 bp) threshold in previous study. Secondly, we included total number of fragments in each 100k bin in the model, as numerous studies have reported the predictive power of cfDNA concentration as a biomarker in cancer diagnosis [3, 26]. Thirdly, we calculated the fragment size distribution of 5 bp bins in each chromosome arm, based on its efficient prediction power from previous study [27]. Finally, we have utilized a more advanced algorithm, stacking ensemble, to learn the fragmentation pattern, as it has successfully improved the prediction performance of the cfDNA model [27–29]. The concept of stacking ensemble is to use unique advantage of different types of ML models, each of which can learn some part of the problem, to generate base models. Another model is then used to learn from the output of these base models for the same problem, leading to the improvement of overall performance. In our model, two different ML algorithms, GBM and SVM, were combined as a stacking ensemble model for the classification.

By applying these strategies, we observed the improvement of the exome-based DELFI score with AUC of 0.896 (95%CI = 0.878 - 0.916) (Figure 2), hereafter called xDELFI. A receiver operator characteristic (ROC) curve indicated the improvement of both sensitivity and specificity (Figure 2 and Supplementary Table S3). By using threshold at 90% specificity, xDELFI showed better sensitivity of 74% in comparison with exomeDELFI which had sensitivity of 69%. We also observed the strong correlation between xDELFI score and the whole genome-based DELFI score (*r* = 0.785) (Figure 2C and Supplementary Table S2) indicating the consistent relationship between both scores.

In order to assess the importance of new features, the model was retrained using a leave-one-feature-out approach. This involves iteratively removing one type of feature at a time before training the model. The contribution of the long fragment feature (>220 bp) and the overall fragment coverage to the prediction performance of xDELFI was the lowest (Supplementary Table S4). On the other hand, the ensemble stacking algorithm had the greatest impact on the prediction performance, followed by FSD.

The model prediction performance on stratifying cancer type revealed that the improvement of xDELFI was observed in almost all cancer types in comparison with the prediction performance of exomeDELFI (Supplementary Table S5-S7). Prediction Performance on the three most abundant cancer types in the dataset was comparable between DELFI and xDELFI model. At 95% specificity, DELFI sensitivity performance on liver cancer (n=93), lung cancer (n=87), and breast cancer (n=60) are 88%, 76% and 57%, respectively, while xDELFI are 87%, 75%, and 53%, respectively. Similarly, the performance on the stratification of cancer stage showed that the improvement of xDELFI was observed in all cancer stages (Supplementary Figure S4 and Table S8). Especially at the most difficult stage I, the exomeDELFI achieved only 53% of sensitivity score at 90% specificity threshold, while xDELFI was able to achieve 60% of the sensitivity score.

To further enhance the sensitivity of cancer detection, we conducted an evaluation of the potential benefits of combining DELFI scores with mutation detection approach. To evaluate this, we analyzed mutation data from a dataset comprising 125 cancer samples [30], and initially found that the targeted mutation approach had a sensitivity of 0.656 for cancer detection. When the exomeDELFI score was combined with the mutation data using the “or” condition, we noticed a significant enhancement in sensitivity from 0.424 to 0.744 (Table 2 and Supplementary Table S1). Additionally, the use the xDELFI score together with the mutation data improved the sensitivity from 0.576 to 0.808. These findings are noteworthy because the standard sequencing coverage of WES is sufficient to detect mutations [31]. Thus, it is possible to calculate the xDELFI score and obtain the mutation profile from WES data, which can be used together to predict cancer patients with higher accuracy than the DELFI approach alone.

**Table 2.** Detection of 125 cancer patients using different DELFI models at 95%Specificity threshold and targeted mutation cfDNA approach.

| Method | TP | FN | Sensitivity |
| --- | --- | --- | --- |
| Mutations | 82 | 43 | 0.656 |
| DELFI | 93 | 32 | 0.744 |
| exomeDELFI | 53 | 72 | 0.424 |
| xDELFI | 72 | 53 | 0.576 |
| DELFI + Mutations | 108 | 17 | 0.864 |
| exomeDELFI+ Mutations | 93 | 32 | 0.744 |
| xDELFI+ Mutations | 101 | 24 | 0.808 |

### xDELFI can predict tissue of origin

One potential application of cfDNA fragmentation profiles is the ability to indicate the tissue of origin. We developed a multiclass machine learning model that can classify the tissue of origin for eight types of cancer, including bile duct, breast, colorectal, gastric, liver, lung, ovarian, and pancreatic cancers. It is important to note that we excluded cancer types with a small sample size (<20) from the model. The performance of the multiclass xDELFI model was comparable to that of the multiclass exome DELFI model, with a mean balanced accuracy of 0.676 (95% CI= 0.598 – 0.754) and 0.666 (95% CI= 0.596 – 0.736), respectively. There was no significant difference between the two models, as determined by a one-sample t-test (*P* = 0.067) (Table 3). However, xDELFI showed greater mean prediction sensitivity with a marginal significant difference (One sample t-test *P* = 0.054) than the exomeDELFI model, although the mean prediction specificity was not different. As expected, the DELFI model outperformed both xDELFI and exome DELFI in all evaluation metrics (One sample t-test *P* < 0.001). The performance of all models was higher than that of random class assignments (Binomial test *P* < 0.001). Assessment of the prediction performance of each class showed that all the models had high specificity but lower sensitivity. Notably, liver, colorectal, and lung cancers had the highest prediction accuracy in all models, likely due to their larger sample sizes, highlighting the importance of sample size in multiclass models (Supplementary Table S9). These findings suggest that the DELFI score based on whole-exome regions can also be used to predict the tissue of origin.

**Table 3.**
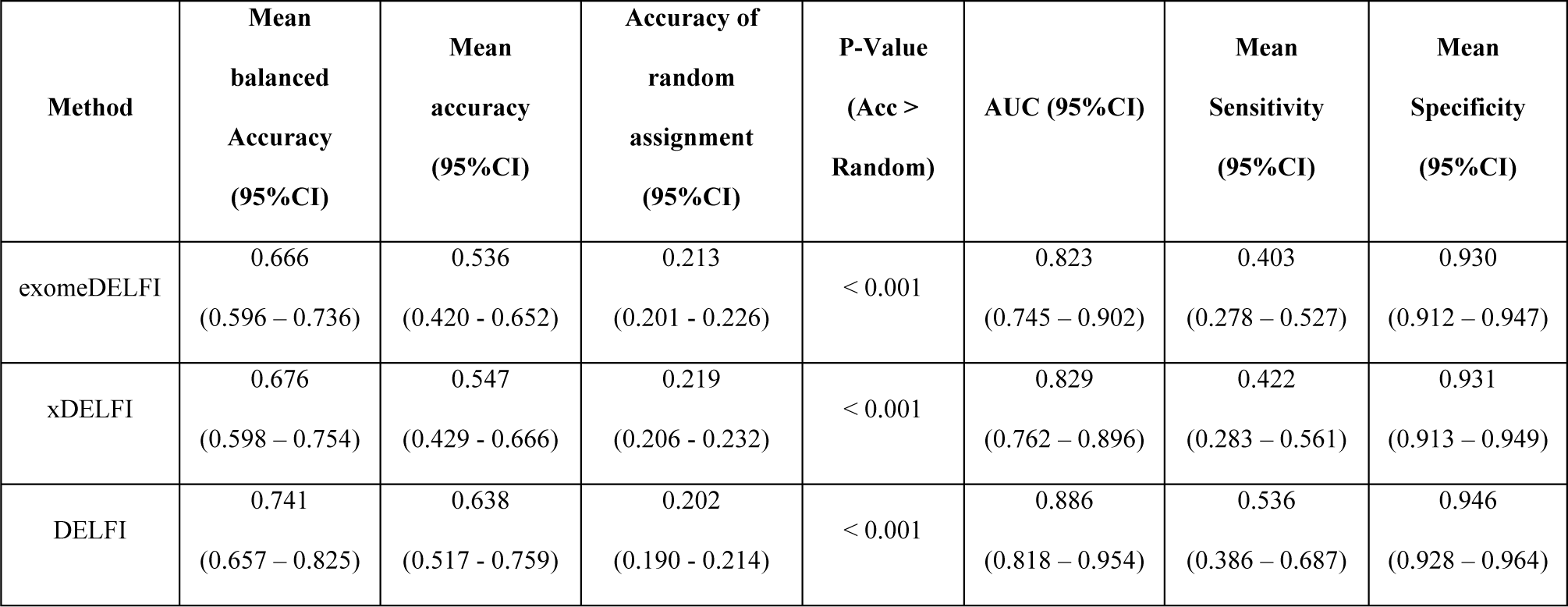
Overall prediction performance of different cfDNA fragmentation models, DELFI, exomeDELFI, and xDELFI on tissue of origin classification.

## Discussion

In our study, we have shown that it is feasible to develop a DELFI model using exome regions. This approach generally offers a significant reduction in sequencing cost, while maintaining a reasonable prediction performance. Interestingly, we observed a notable distinction in the feature profile between the WGS-based DELFI and exome-based DELFI, despite the prediction trend being similar. This finding suggests the potential for novel features that can be incorporated into the existing DELFI model to improve prediction accuracy.

We demonstrated that implementing a new feature extraction scheme and utilizing more advanced algorithms can significantly improve the accuracy of the exome-based DELFI model. While short (100-150 bp) and long (151-220 bp) cfDNA fragment coverage within each window were the key features utilized in the DELFI model, the model performance can be enhanced by incorporating new fragmentomic features such as fragmentation size coverage (FSC), fragmentation size distribution (FSD), and employing ensemble stacking algorithms [28]. With these strategies, we can also improve exome-based DELFI performance in xDELFI. Further analysis using leave-one-feature-out approach to determine feature importance revealed that the long fragment feature (>220 bp) had the lowest contribution to the prediction performance of xDELFI. In contrast, the ensemble stacking algorithm was found to have the most contribution to the prediction performance, followed by FSD and overall coverage. These findings suggest that exome-based DELFI performance can be improved without incurring additional sequencing costs. According to suggestion in Liu’s 2021 review [32], new feature extraction strategies should be developed to further enhance DELFI performance, for example, three-dimensional chromatin organization [33], and cfDNA-accessibility score near the transcription factor-binding sites [34]. Additionally, several studies have demonstrated a correlation between cfDNA-fragmentation patterns at the transcription start site (TSS) and gene expression [14, 35, 36]. Thus, incorporating WES plus untranslated region (UTR) could potentially provide unique features and improve the prediction performance of DELFI.

The DELFI, exomeDELFI, and xDELFI approaches produce scores that are highly correlated with each other. However, the fragmentation patterns observed in whole genome and whole exome are different suggesting that the fragmentation pattern is not uniformly distributed across all regions. The exonic regions, which are regions of DNA that encode proteins, holds specific fragmentation pattern different from the pattern of non-exonic regions, and can influence the prediction performance of the model. These findings are consistent with previous research showing that exonic and intronic regions have different expression and chromatin patterns [37].

The performance of the DELFI score has been shown to be enhanced when combined with mutation detection results [15]. However, implementing two sequencing services, one for DELFI score and the other for targeted mutation detection, can be expensive and may not be feasible in practical scenarios [38]. Thus, calculating xDELFI score from WES technology is an alternative approach with comparable accuracy to DELFI score at a more affordable cost. WES can provide mutation information that can be utilized in together with the xDELFI score to improve prediction sensitivity up to 81%. However, it is important to conduct a systemic study to validate the actual performance of this approach with actual WES data, which may subject to significant bias caused by exome capture probs [39]. Therefore, a new method for correcting prob-bias is required. In summary, our findings suggest that the use of exome regions is a viable alternative for developing the DELFI score, given its reasonable accuracy and affordable cost.

## Data Availability Statement

All code and data generated or analyzed during this study are deposited in Github: https://github.com/asangphukieo/xDELFI.

## Acknowledgments

This work was supported by Faculty of Medicine Research Fund, Chiang Mai University under award number 031-2566. This work was supported by the Google Cloud Research Credits program with the award GCP19980904.

## Author Contributions Statement

A.S. and P.C. conceived of the presented idea. A.S. developed the theory, designed the model and the computational framework, and analysed the data. P.N. and P.T. verified the analytical methods. P.C. helped supervise the project. A.S. wrote the manuscript in consultation with P.C., P.T. and P.N. All authors discussed the results and contributed to the final manuscript.

## Supplementary data

**Figure S1** Proportion of cancer patients in each type and healthy individuals in the dataset

**Figure S2** Correlation between sequence coverage from original article and from this study

**Figure S3** Correlation between DELFI score reported in 2019 and DELFI score reported in this study

Figure S4 Violin plot of DELFI, exomeDELFI, and xDELFI score in each cancer stage

**Table S1** DELFI score, exomeDELFI score and xDELFI score of 721 samples

**Table S2** Pearson’s correlation coefficient of different DELFI scores

**Table S3** Prediction performance of different cfDNA fragmentation models, DELFI, exomeDELFI, and xDELFI

**Table S4** Leave-one-out prediction performance on feature type of xDELFI model at 95% specificity threshold

**Table S5** DELFI prediction performance of 721 samples on the stratification of cancer types

**Table S6** ExomeDELFI prediction performance of 721 samples on the stratification of cancer types

**Table S7** xDELFI prediction performance of 721 samples on the stratification of cancer types

**Table S8** Prediction performance of different cfDNA fragmentation models, DELFI, exomeDELFI, and xDELFI on the stratification of cancer stage

**Table S9** Prediction performance of each class of different cfDNA fragmentation models, DELFI, exomeDELFI, and xDELFI on tissue of origin classification

